# SARS-CoV-2 Infections Among Children in the Biospecimens from Respiratory Virus-Exposed Kids (BRAVE Kids) Study

**DOI:** 10.1101/2020.08.18.20166835

**Authors:** Jillian H. Hurst, Sarah M. Heston, Hailey N. Chambers, Hannah M. Cunningham, Meghan J. Price, Liliana Suarez, Carter G. Crew, Shree Bose, Jhoanna N. Aquino, Stuart T. Carr, S. Michelle Griffin, Stephanie H. Smith, Kirsten Jenkins, Trevor S. Pfeiffer, Javier Rodriguez, C. Todd DeMarco, Nicole A. De Naeyer, Thaddeus C. Gurley, Raul Louzao, Coleen K. Cunningham, William J. Steinbach, Thomas N. Denny, Debra J. Lugo, M. Anthony Moody, Sallie R. Permar, Alexandre T. Rotta, Nicholas A. Turner, Emmanuel B. Walter, Christopher W. Woods, Matthew S. Kelly

## Abstract

**BACKGROUND:** Children with SARS-CoV-2 infection typically have mild symptoms that do not require medical attention, leaving a gap in our understanding of the spectrum of illnesses that the virus causes in children.

**METHODS:** We conducted a prospective cohort study of children and adolescents (<21 years of age) with a SARS-CoV-2-infected close contact. We collected nasopharyngeal or nasal swabs at enrollment and tested for SARS-CoV-2 using a real-time PCR assay.

**RESULTS:** Of 382 children, 289 (76%) were SARS-CoV-2-infected. SARS-CoV-2-infected children were more likely to be Hispanic (p<0.0001), less likely to have a history of asthma (p=0.009), and more likely to have an infected sibling contact (p=0.0007) than uninfected children. Children ages 6-13 years were frequently asymptomatic (38%) and had respiratory symptoms less often than younger children (30% vs. 49%; p=0.008) or adolescents (30% vs. 59%; p<0.0001). Compared to children ages 6-13 years, adolescents more frequently reported influenza-like (61% vs. 39%; p=0.002), gastrointestinal (26% vs. 9%; p=0.003), and sensory symptoms (43% vs. 9%; p<0.0001), and had more prolonged illnesses [median (IQR) duration: 7 (4, 12) vs. 4 (3, 8) days; p=0.004]. Despite the age-related variability in symptoms, we found no differences in nasopharyngeal viral load by age or between symptomatic and asymptomatic children.

**CONCLUSIONS:** Hispanic ethnicity and an infected sibling close contact are associated with increased SARS-CoV-2 infection risk among children, while a history of asthma is associated with decreased risk. Age-related differences in the clinical manifestations of SARS-CoV-2 infection must be considered when evaluating children for COVID-19 and in developing screening strategies for schools and childcare settings.

## INTRODUCTION

Severe acute respiratory syndrome coronavirus 2 (SARS-CoV-2) has been responsible for more than 20 million infections and 750,000 deaths as of August 2020. Current epidemiological data suggest children are less susceptible to SARS-CoV-2 infection than adults. Population screening in Iceland found SARS-CoV-2 was detected at a lower rate among children <10 years of age compared with adolescents and adults (6.7% vs. 13.7%).^1^ Further, recent mathematical modeling from Asia and Europe estimated susceptibility of individuals <20 years of age to the virus was approximately half that of older adults.^2^ Finally, in a household transmission study, the secondary attack rate was lower among children less than 20 years of age (5%) than among adults 20-59 years of age (15%) or 60 years of age or older (18%).^3^ The extent to which these findings reflect differences in SARS-CoV-2 exposures among adults and children or age-related biological differences in SARS-CoV-2 susceptibility is unknown. Thus far, few factors that influence infection risk among SARS-CoV-2-exposed children have been identified.

Children infected with SARS-CoV-2 generally have milder illnesses than adults. In a recent meta-analysis of data from 371 children <18 years of age, fever (51%) and cough (37%) were the most frequently reported symptoms, while 17% of children were asymptomatic.^4^ To date, studies describing the clinical characteristics of SARS-CoV-2 infections among children have been limited by cross-sectional designs, small sample sizes, or inclusion of only hospitalized or symptomatic children.^5-9^ Given that a small minority of children with SARS-CoV-2 infection require hospitalization, the spectrum of illnesses caused by SARS-CoV-2 in children has not been well characterized. Such data are critical for providers evaluating children with possible coronavirus disease 2019 (COVID-19) and for the development of effective screening strategies for children to attend schools and other congregate childcare settings.

We describe risk factors, clinical manifestations, and nasopharyngeal viral loads of SARS-CoV-2 infection among 382 children and adolescents living within the catchment area of a health system in central North Carolina, constituting the largest non-hospitalized pediatric cohort described to date.

## METHODS

### Study Design

The Duke Biospecimens from RespirAtory Virus-Exposed Kids (BRAVE Kids) study is a prospective cohort study of children and adolescents with confirmed SARS-CoV-2 infection or close contact with an individual with confirmed SARS-CoV-2 infection. This study is being conducted within the Duke University Health System (DUHS) in Raleigh-Durham, North Carolina. The DUHS is a large, integrated health system consisting of three hospitals and over 100 outpatient clinics. This study was approved by the DUHS Institutional Review Board.

### Study Participants

Eligible participants were <21 years of age and had close contact with an individual with laboratory-confirmed SARS-CoV-2 infection. Participants were identified either through presentation to the health system themselves or through presentation of a close contact with laboratory-confirmed SARS-CoV-2 infection. We defined close contact as an unprotected exposure within 6 feet to a confirmed case between 2 days before and 7 days after symptom onset or laboratory confirmation of SARS-CoV-2 infection in asymptomatic contacts. Close contacts included, but were not limited to, parents, siblings, other caregivers, partners, and relatives. Informed consent was obtained from study participants or their legal guardians; assent was obtained for children 8-17 years of age. Written consent was provided using an electronic consent document. We obtained a waiver of documentation for participants who did not have an email address or were unable to complete the electronic consent document.

### Study Procedures

We collected exposure, sociodemographic, and clinical data at enrollment through review of electronic medical records and a directed caregiver questionnaire conducted by telephone. We recorded symptoms occurring up to 14 days prior to enrollment. Research staff conducted follow-up questionnaires by phone for all participants 7 days after study enrollment to document new symptoms and healthcare encounters. For participants with ongoing symptoms 7 days after study enrollment, additional questionnaires were administered 14 and 28 days after enrollment, or until the participant reported complete symptom resolution. We recorded the results of SARS-CoV-2 testing performed for clinical care. Research staff collected nasopharyngeal swabs from participants who consented to a home visit. Participants who declined a home visit received a kit for self-collection of a mid-turbinate nasal swab. Nasopharyngeal and nasal samples were collected with nylon flocked swabs (Copan Italia, Brescia, Italy) into RNAProtect (Qiagen, Hilden, Germany).

### Viral Load Assay

SARS-CoV-2 RNA copies per milliliter (copies/mL) was determined by a two-step real-time quantitative PCR assay developed in the Clinical Laboratory Improvement Amendments-certified Immunology and Virology Quality Assessment Center at the Duke Human Vaccine Institute. DSP Virus/Pathogen Midi Kits (Qiagen, Hilden, Germany) were used to extract viral RNA on a QIAsymphony SP automated sample preparation platform. A reverse primer specific to the SARS-CoV-2 envelope gene was annealed to the extracted RNA and reverse transcribed into cDNA using SuperScript III Reverse Transcriptase and RNaseOut (Thermo Fisher Scientific, Waltham, MA). cDNA was treated with RNase H and then added to a custom 4x TaqMan Gene Expression Master Mix (Applied Biosystems, Foster City, CA) containing envelope gene-specific primers and a fluorescently labeled hydrolysis probe; quantitative PCR was carried out on a QuantStudio 3 Real-Time PCR system (Thermo Fisher Scientific, Waltham, MA).^10^ SARS-CoV-2 RNA copies per reaction were interpolated using quantification cycle data and a serial dilution of a highly characterized custom DNA plasmid containing the SARS-CoV-2 envelope gene sequence. The limit of quantification was 62 RNA copies/mL of sample as determined by an extensive validation process consistent for use in a clinical setting.

### Data Analysis

We described characteristics of the study population by SARS-CoV-2 infection status using frequencies and percentages for categorical variables, and medians and interquartile ranges (IQR) for continuous variables. We used chi-square or Fisher’s exact tests for categorical variables and Wilcoxon rank-sum tests or ANOVA for continuous variables to compare the characteristics of SARS-CoV-2-infected and uninfected children and to evaluate age-related differences in symptoms among SARS-CoV-2-infected children. We compared nasopharyngeal SARS-CoV-2 viral loads (measured as log_10_ copies/mL) by age, illness characteristics, and timing of sample collection relative to symptom onset using ANOVA or linear regression. We used a quantile-quantile plot to verify normality of the nasopharyngeal viral load data. Study data were managed using REDCap electronic data capture tools hosted at Duke University.^11^ Analyses were performed using R version 3.6.1.^12^

## RESULTS

### Patient Characteristics

Among the 382 children enrolled between April 7 and July 16, 2020 (**Figure 1**), median (IQR) age was 9.8 (4.9, 15.9) years, 205 (54%) children were female, and 309 (81%) subjects were of Hispanic ethnicity. Most children were healthy, with the most commonly identified comorbidities being obesity (body mass index ≥95^th^ percentile for age; 29%) and a history of asthma (9%). Two hundred eighty-nine (76%) children were SARS-CoV-2-infected and 93 (24%) were SARS-CoV-2-uninfected (**Table 1**). A history of asthma was less common in SARS-CoV-2-infected children than in uninfected children (7% vs. 16%; p=0.009). SARS-CoV-2-infected children were more likely to be of Hispanic ethnicity (88% vs. 58%; p<0.0001) and to have an infected sibling contact than uninfected children (50% vs. 29%; p=0.0007). Of 144 SARS-CoV-2-infected children with an infected sibling, 45 of 144 (31%) did not have any identified adult close contacts with confirmed SARS-CoV-2 infection. Among these 45 children, median (IQR) age of the infected sibling contacts was 12.4 (8.4, 16.3) years.

**Figure 1.**
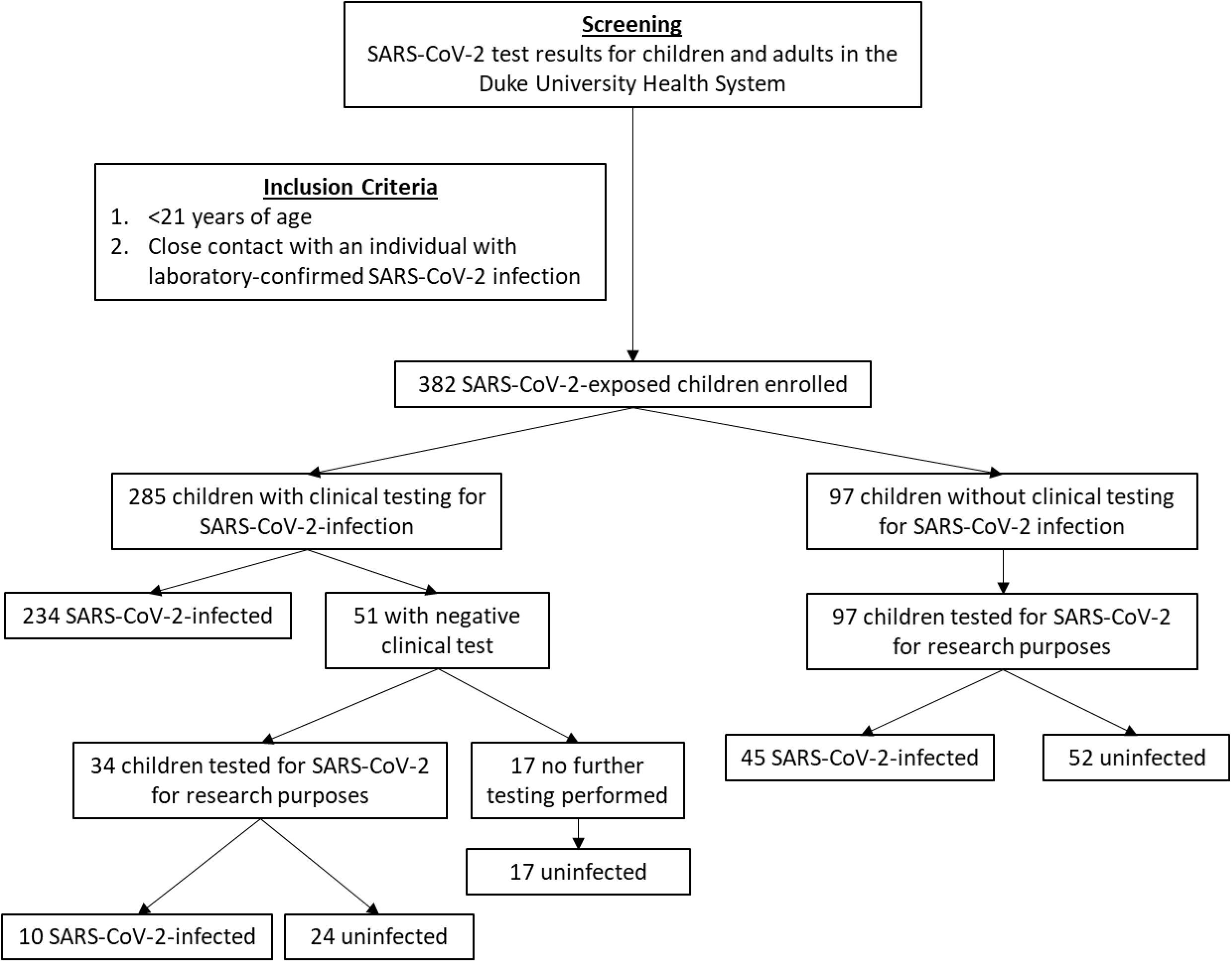
Flowchart of enrollment and determination of SARS-CoV-2 infection status in the study population

**Table 1.** Characteristics of the study population

|  |  | **Total**<br>**(n=382)** |  | **SARS-CoV-2-Infected**<br>**(n=289)** |  | **SARS-CoV-2-Uninfected**<br>**(n=93)** |  | ***P*** |
| --- | --- | --- | --- | --- | --- | --- | --- | --- |
|  |  | N (or median) | % (or IQR) | N (or median) | % (or IQR) | N (or median) | % (or IQR) |  |
| Age, years |  | 9.8 | (4.9, 15.9) | 10.5 | (4.9, 16.4) | 8.9 | (5.0, 14.7) | 0.36 |
| Sex |  |  |  |  |  |  |  | 0.92 |
|  | Female | 205 | 54% | 156 | 54% | 49 | 53% |  |
|  | Male | 177 | 46% | 133 | 46% | 44 | 47% |  |
| Race |  |  |  |  |  |  |  | <0.0001 |
|  | Black or African-American | 26 | 7% | 17 | 6% | 9 | 10% |  |
|  | Latino or Hispanic-American | 309 | 81% | 255 | 88% | 54 | 58% |  |
|  | Non-Hispanic white | 45 | 12% | 16 | 6% | 29 | 31% |  |
|  | Other | 2 | <1% | 1 | <1% | 1 | 1% |  |
| Number of household members |  | 5 | (4, 6) | 5 | (4, 6) | 5 | (4, 6) | 0.77 |
| Close contacts with SARS-CoV-2 |  |  |  |  |  |  |  |  |
|  | Parent | 217 | 57% | 157 | 54% | 60 | 65% | 0.11 |
|  | Sibling | 171 | 45% | 144 | 50% | 27 | 29% | 0.0007 |
|  | Other | 103 | 27% | 76 | 26% | 27 | 29% | 0.70 |
| Comorbidities |  |  |  |  |  |  |  |  |
|  | Provider-diagnosed asthma | 34 | 9% | 19 | 7% | 15 | 16% | 0.009 |
| | Obesity (BMI ≥ 95<sup>th</sup> percentile for age) | 109 | 29% | 87 | 30% | 22 | 24% | 0.29 |

IQR, interquartile range; BMI, body mass index

### Symptoms of SARS-CoV-2 Infection

One or more symptoms were reported at enrollment or in follow-up by 204 (71%) subjects with confirmed SARS-CoV-2 infection (**Table 2**). The most commonly reported symptoms were fever (42%), cough (34%), and headache (26%). The median (IQR) duration of symptoms was 5 (3-10) days; 90% of symptomatic children reported full symptom resolution within 15 days. The clinical manifestations of SARS-CoV-2 infection varied by age (**Figure 2**). Symptoms were reported in 75% of children ages 0-5 years, 62% of children ages 6-13 years, and 76% of adolescents age 14-20 years (p=0.048). Children 6-13 years of age reported respiratory symptoms less often than younger children (30% vs. 49%; p=0.008) and adolescents 14-20 years of age (30% vs. 60%; p<0.0001). Compared to children 6-13 years of age, adolescents 14-20 years of age also more frequently reported influenza-like (61% vs. 39%; p=0.002), gastrointestinal (26% vs. 9%; p=0.003), and sensory symptoms (43% vs. 9%; p<0.0001). Adolescents had more prolonged illnesses than either children ages 0-5 years [median (IQR) duration: 7 (4, 12) vs. 4 (3, 7.75) days; p=0.002] or children ages 6-13 years median (IQR) duration: 7 (4, 12) vs. 4 (3, 8) days; p=0.004]. One infant with a prior history of severe bronchiolitis required hospitalization for respiratory distress and was given remdesivir.

**Figure 2.**
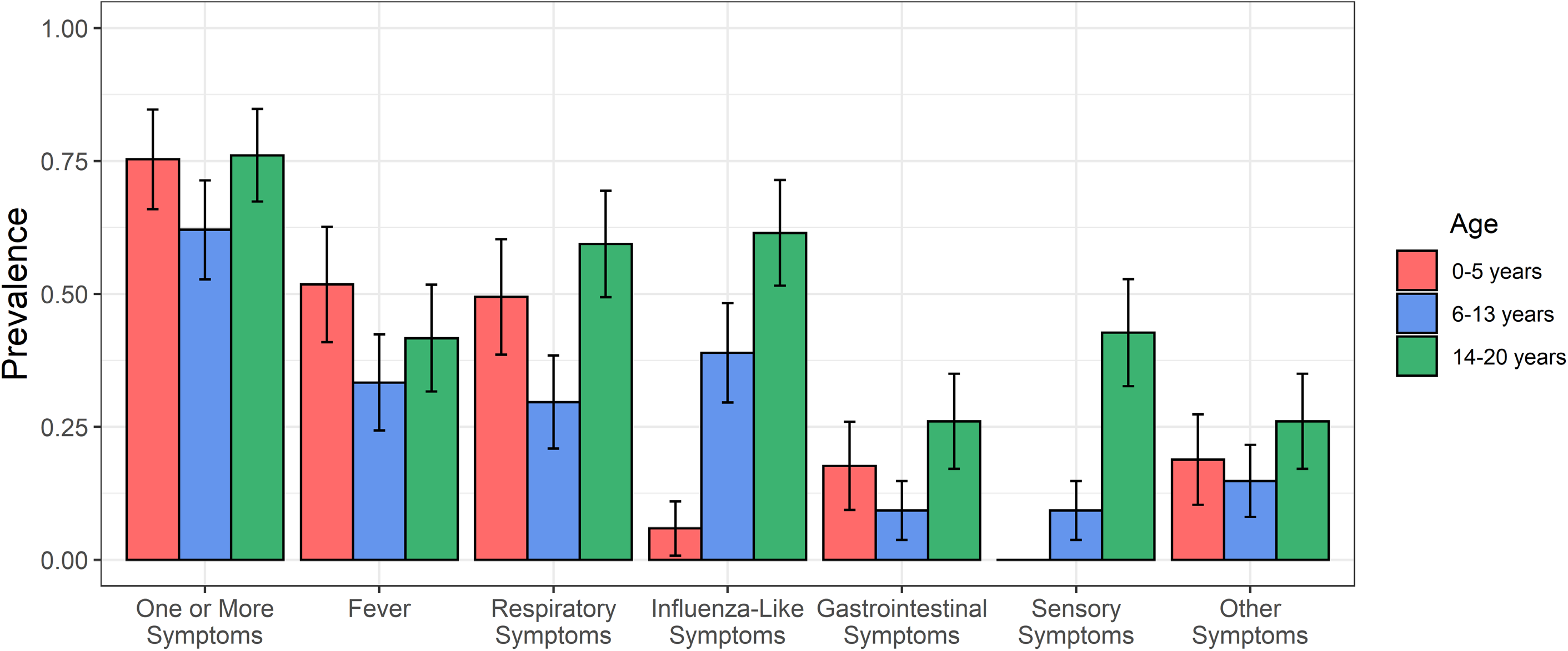
Prevalence of reported symptom complexes in 289 SARS-CoV-2-infected children by age. Age was categorized into three groups (0-5 years, 6-13 years, and 14-20 years), and the prevalence of specific symptom complexes are reported for children in each age group. Symptom complexes include respiratory symptoms (cough, difficulty breathing, nasal congestion, or rhinorrhea), influenza-like symptoms (headache, myalgias, or pharyngitis), gastrointestinal symptoms (abdominal pain, diarrhea, or vomiting), and sensory symptoms (anosmia or dysgeusia). Error bars correspond to the 95% confidence interval for each symptom complex in each age group.

**Table 2.** Clinical manifestations of SARS-CoV-2 infection among children and adolescents

|  | Total<br>(n=289) |  | 0-5 Years<br>(n=85) |  | 6-13 Years<br>(n=108) |  | 14-20 Years<br>(n=96) |  | P |
| --- | --- | --- | --- | --- | --- | --- | --- | --- | --- |
|  | n (%) |  | n (%) |  | n (%) |  | n (%) |  |  |
| Asymptomatic infection | 85 | 29% | 21 | 25% | 41 | 38% | 23 | 24% | 0.048 |
| Symptomatic infection |  |  |  |  |  |  |  |  |  |
| Median (IQR) days of symptoms | 5 | (3, 10) | 4 | (3, 7.75) | 4 | (3, 8) | 7 | (4, 12) | 0.004 |
| Fever | 120 | 42% | 44 | 52% | 36 | 33% | 40 | 42% | 0.04 |
| Respiratory symptoms | 131 | 45% | 42 | 49% | 32 | 30% | 57 | 59% | <0.0001 |
| Cough | 98 | 34% | 26 | 31% | 25 | 23% | 47 | 49% |  |
| Difficulty breathing | 29 | 10% | 7 | 8% | 9 | 8% | 13 | 14% |  |
| Nasal congestion | 34 | 12% | 12 | 14% | 5 | 5% | 17 | 18% |  |
| Rhinorrhea | 31 | 11% | 14 | 16% | 1 | <1% | 16 | 16% |  |
| Influenza-like symptoms | 106 | 37% | 5 | 6% | 42 | 39% | 59 | 61% | <0.0001 |
| Headache | 74 | 26% | 4 | 5% | 23 | 21% | 47 | 49% |  |
| Myalgias | 49 | 17% | 1 | 1% | 18 | 17% | 30 | 31% |  |
| Pharyngitis | 44 | 15% | 1 | 1% | 13 | 12% | 30 | 31% |  |
| Gastrointestinal symptoms | 50 | 17% | 15 | 18% | 10 | 9% | 25 | 26% | 0.007 |
| Abdominal pain | 20 | 7% | 5 | 6% | 6 | 6% | 9 | 9% |  |
| Diarrhea | 30 | 10% | 11 | 13% | 5 | 5% | 14 | 15% |  |
| Vomiting | 20 | 7% | 4 | 5% | 3 | 3% | 13 | 14% |  |
| Sensory symptoms | 51 | 18% | 0 | 0% | 10 | 9% | 41 | 43% | <0.0001 |
| Anosmia | 43 | 15% | 0 | 0% | 10 | 9% | 33 | 34% |  |
| Dysgeusia | 43 | 15% | 0 | 0% | 9 | 8% | 34 | 35% |  |
| Other symptoms | 57 | 20% | 16 | 19% | 16 | 15% | 25 | 26% | 0.13 |
| Arthralgias | 10 | 3% | 2 | 2% | 1 | <1% | 7 | 7% |  |
| Chest pain | 11 | 4% | 0 | 0% | 3 | 3% | 8 | 8% |  |
| Conjunctivitis | 7 | 2% | 2 | 2% | 0 | 0% | 5 | 5% |  |
| Rash | 8 | 3% | 6 | 7% | 0 | 0% | 2 | 2% |  |

IQR, interquartile range

### Nasopharyngeal Viral Loads

We performed quantitative SARS-CoV-2 PCR on nasopharyngeal samples from 260 study participants. SARS-CoV-2 was detected in 179 (69%) samples at a median (IQR) viral load of 4.0 (3.0, 5.6) log copies/mL. We evaluated associations between nasopharyngeal viral load and age, symptoms, and the timing of sample collection relative to symptom onset (**Figure 3**). SARS-CoV-2 viral loads did not differ by age group (p=0.54). Amongst symptomatic children, nasopharyngeal viral loads were highest in the 3 days before and after onset of symptoms and declined with increasing time from symptom onset (p<0.0001). Nasopharyngeal viral loads did not differ in asymptomatic and symtpomatic children of any age [median (IQR): 3.7 (2.6, 6.5) vs. 4.1 (3.0, 5.4) log copies/mL; p=0.53]; similarly, we found no association between viral load and the presence of fever, respiratory symptoms, or other reported symptom complexes.

**Figure 3.**
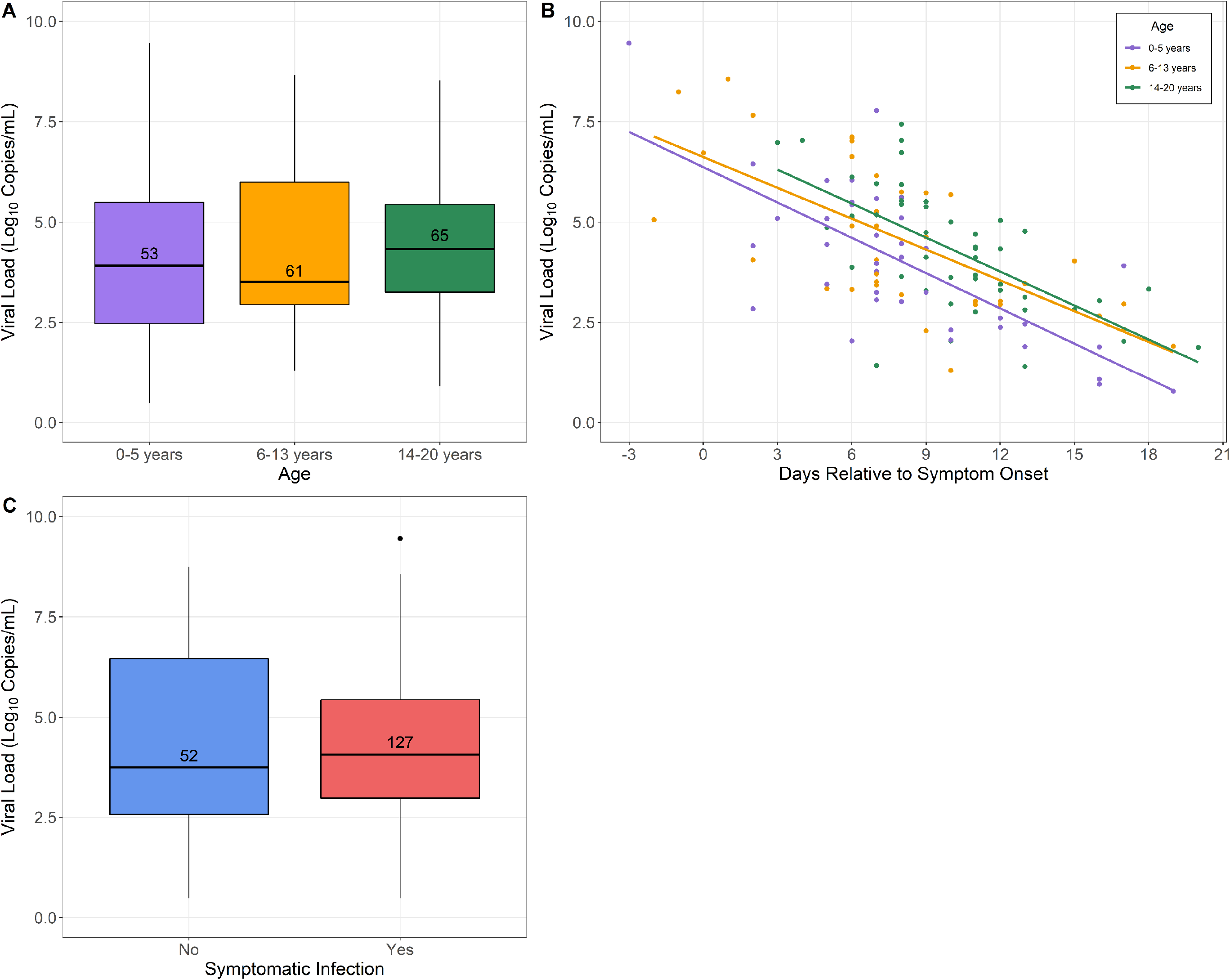
Evaluation of nasopharyngeal SARS-CoV-2 viral load among 178 SARS-CoV-2-infected children by age, symptoms, and timing of sample collection relative to symptom onset. Panel A shows viral loads among SARS-CoV-2-infected children by age group; no difference in viral load was seen with respect to age (p=0.54). Panel B shows viral loads in symptomatic SARS-CoV-2-infected children relative to the timing of symptom onset (days -3 to 20). SARS-CoV-2 viral loads were highest in the 3 days before and after symptom onset [median (IQR): 6.6 (4.9, 7.8) log copies/mL] and declined with increasing time from symptom onset (p<0.0001). Adjusting for the timing of sample collection relative to symptom onset, there were no differences in nasopharyngeal viral load by age group (0-5 years vs. 14-20 years, p=0.10; 6-13 years vs. 14-20 years, p=0.53). Panel C shows viral loads among SARS-CoV-2-infected children who reported one or more symptoms and children who reported no symptoms; viral loads were similar among asymptomatic children and children with symptomatic COVID-19 [median (IQR): 3.7 (2.6, 6.5) vs. 4.1 (3.0, 5.4) log copies/mL; p=0.53].

## DISCUSSION

We describe the clinical and epidemiological characteristics of 382 children and adolescents who had close contact with a SARS-CoV-2-infected individual. We found that Hispanic ethnicity and a SARS-CoV-2-infected sibling were risk factors for SARS-CoV-2 infection, while a history of asthma was associated with a decreased infection risk. We also report that the characteristics and duration of illnesses among SARS-CoV-2-infected children vary by age. Finally, we demonstrate that nasopharyngeal SARS-CoV-2 viral loads do not differ by age or between symptomatic and asymptomatic children, and decrease sharply after symptom onset in children and adolescents.

More than 80% of children in our cohort were Hispanic, and Hispanic ethnicity was associated with an increased risk of SARS-CoV-2 infection. Individuals of Hispanic ethnicity account for 59-62% of all SARS-CoV-2 cases reported in the study catchment area.^13^ Analyses of SARS-CoV-2 infections in New York and Houston identified similar racial and ethnic disparities in infection risk.^14,15^ We also found that having an infected sibling was a risk factor for SARS-CoV-2 infection. Early studies suggested that children transmit SARS-CoV-2 less effectively than adults, but evidence for efficient transmission from children has been accumulating.^9,16-18^ Further, there have been increasing reports of infections among children as schools, camps, and other childcare facilities reopen in the United States and other countries.^17,19,20^ Nearly one-third of the SARS-CoV-2-infected children who had an infected sibling in our cohort did not have any other known infected close contacts, suggesting probable child-to-child transmission within these households.

Our findings suggest that asthma is associated with a lower susceptibility to SARS-CoV-2 infection among children. Though many viral respiratory infections are associated with asthma exacerbations, a recent study of adults hospitalized with SARS-CoV-2 pneumonia found no difference in disease severity between asthmatic and non-asthmatic patients.^21^ Several prior studies reported that individuals with asthma are underrepresented in cohorts of patients with COVID-19.^22-24^ In a study of 1590 individuals hospitalized for COVID-19 in China, not a single patient had a history of provider-diagnosed asthma.^24^ These observations have led to speculation that asthma may lower SARS-CoV-2 susceptibility, or alternatively protect from severe COVID-19, by promoting a Th2-dominant immune response or through reduced expression of the SARS-CoV-2 receptor (ACE2).^25^

Consistent with prior reports, we found that the majority of SARS-CoV-2-infected children had mild illnesses, with only a single subject requiring hospitalization for COVID-19. Moreover, symptoms reported by children in our cohort were broadly similar to those seen in other pediatric studies.^4,5,7,8^ Among 291 SARS-CoV-2-infected children with symptom data reported to the Centers for Disease Control and Prevention (CDC), fever, cough, and headache were most commonly reported.^26^ Similar to recent studies of SARS-CoV-2-infected adults, gastrointestinal, and sensory symptoms (anosmia or dysgeusia) were relatively common in our cohort.^27,28^ However, we found that the clinical manifestations of SARS-CoV-2 infection among children and adolescents varied markedly by age. Approximately 75% of children <6 years of age had symptomatic infection most frequently characterized by fever or cough. By comparison, only 62% of children 6-13 years of age were symptomatic, and less than one-third of these children reported respiratory symptoms. Symptoms reported by SARS-CoV-2-infected adolescents were generally similar to those described in adults, with high prevalence of respiratory, influenza-like, gastrointestinal, and sensory symptoms.^29^ Illness duration among SARS-CoV-2-infected children in our cohort increased with age. Symptom duration among children <13 years of age was shorter than among older adolescents, and all age groups had shorter illness durations than have generally been reported in adults.^30,31^ In a study of 270 outpatient SARS-CoV-2-infected adults in the United States, 35% of adults reported not having returned to their usual state of health 14 to 21 days after SARS-CoV-2 testing.^30^

Recent studies evaluating associations between age and nasopharyngeal viral load reported conflicting results. Among 145 children and adults with symptomatic SARS-CoV-2 infection in Chicago, higher amounts of viral nucleic acid were detected in samples from 46 children <5 years of age than from 51 older children and 48 adults.^32^ This study used cycle threshold (Ct) values from a PCR assay that has been approved for clinical use, but has not been calibrated for quantitation.^32^ A study conducted in Switzerland showed no difference in nasopharyngeal viral loads between 53 children <11 years of age and adults.^33^ In this largest pediatric cohort reported to date, we found no association between age and nasopharyngeal SARS-CoV-2 viral load among children and adolescents <21 years of age. Our findings indicate that, despite marked age-related differences in the clinical manifestations of SARS-CoV-2 infection, viral load in the upper respiratory tract is similar across the age spectrum. Conflicting data have also been reported with regard to associations between nasopharyngeal viral load and illness severity.^34-36^ A higher nasopharyngeal viral load predicted a shorter duration of illness among adults presenting for emergency care, while a higher viral load was associated with an increased risk of intubation in hospitalized adults.^35,36^ Moreover, a prior study suggested that asymptomatic patients have viral loads that approximate those of patients with symptomatic COVID-19.^37^ In our pediatric cohort, nasopharyngeal viral loads were similar across age groups and did not differ based on symptoms. Finally, as previously described in adults, we found a strong association between the timing of symptom onset and nasopharyngeal viral load, with the highest viral loads among children and adolescents observed around the time of symptom onset.^38^

Our study has several limitations. First, study recruitment was influenced by local SARS-CoV-2 testing availability and guidelines, which changed substantially during the study period and may differ from other areas. Given our study design and the relatively high rate of asymptomatic infection among children in our cohort, we were unable to determine the direction of SARS-CoV-2 transmission within households. Nearly one-third (30%) of children in our cohort were tested for SARS-CoV-2 infection at only a single time point, and some children who ultimately developed SARS-CoV-2 infection may have been misclassified as uninfected because of the timing of sample collection. The prevalence of influenza-like and sensory symptoms should be interpreted with caution in children <5 years of age, given that many children in this age group are unable to verbalize these symptoms. Further, viral loads from nasopharyngeal swabs are likely affected by sampling technique. Finally, analyses were limited to detection of viral nucleic acid, although a prior study reported a close correlation between viral load and infectious virus in symptomatic neonates, children, and adolescents.^39^

In summary, we identify risk factors for SARS-CoV-2 infection among children and present further evidence of probable child-to-child transmission within household settings. Moreover, we demonstrate that the clinical manifestations of SARS-CoV-2 infection among children and adolescents are dependent on age. Finally, we show that children and adolescents with SARS-CoV-2 infection have similar nasopharyngeal viral loads. Future studies are needed to elucidate the biological and immunological factors that account for the age-related differences in infection susceptibility and illness characteristics among children.

## Declarations

SRP consults for cytomegalovirus vaccine programs at Merck, Sanofi, Moderna, and Pfizer, and receives support for research from Moderna and Merck. EBW is an investigator for clinical trials funded by Pfizer and Moderna. All other authors have no conflicts of interest to declare.

## Data Availability

The data that support the findings of this study are available from the corresponding author, MSK, upon reasonable request.

## Acknowledgments

We offer sincere gratitude to the children and families who participated in this research.

